# Drug-Drug interactions between COVID-19 treatments and antipsychotics drugs: integrated evidence from 4 databases and a systematic review

**DOI:** 10.1101/2020.06.04.20122416

**Authors:** Plasencia-García Beatriz Oda, Rodriguez-Menendez Gonzalo, Rico-Rangel María Isabel, Rubio-Garcia Ana, Torelló-Iserte Jaime, Crespo-Facorro Benedicto

## Abstract

**Relevance:** Management of symptoms like anxiety, delirium and agitation cannot be neglected in COVID-19 patients. Antipsychotics are usually used for the pharmacological management of delirium, and confusion and behavioral disturbances. The selection of concomitant COVID-19 medications and antipsychotics should be evidence-based and closely monitored

**Objective:** To systematically review evidence-based available on drug-drug interactions between COVID-19 treatments and antipsychotics.

**Evidence Review:** Three databases were consulted: (a) Lexicomp® Drug Interactions, (b) Micromedex® Solutions Drugs Interactions, (c) Liverpool © Drug Interaction Group for COVID-19 therapies. To acquire more information on QT prolongation and TdP, the CredibleMeds® QTDrugs List was searched. Based on the information collected, the authors made a recommendation agreed to by consensus. In addition, a systematic review was conducted to find the clinical outcomes of drug-drug interactions between COVID-19 treatments and antipsychotics

**Results:** The main interaction between COVID-19 drugs and antipsychotics are the risk of QT prolongation and TdP, and CYP interactions. Remdesivir, favipiravir, baricinitib, and anakinra can be used concomitantly with antipsychotics with no risk of drug-drug interaction (except for hematological risk with clozapine and baricinitib). Tocilizumab is rather safe to use in combination with antipsychotics, although it can restore the activity of CYP3A4 and therefore its substrate metabolism may increase. The most demanding COVID-19 treatments for co-administration with antipsychotics are chloroquine, hydroxychloroquine, azithromycin (all prolong QT interval) and lopinavir / ritonavir (CYP interaction and risk of QT prolongation).

**Conclusions:** We urge to development of evidence-based guidelines that can help clinicians decide the safest treatment combination and monitoring necessary for each particular patient. The selection of concomitant COVID-19 medications and antipsychotics should be evidence-based and closely monitored.

## INTRODUCTION

The number of people affected by coronavirus disease 2019 (COVID-19) worldwide is over 4.5M and rapidly increasing, and the importance of knowledge of symptom management and treatment for general practitioners and other clinicians must be stressed. Although the key symptoms are cough, fever, and breathlessness, anxiety, delirium, and agitation are also present in a large number of patients^1^. Up to 20–30% of COVID-19 patients will have with or develop delirium or changes in the mental health during their hospitalizations, and these numbers are as high as 60–70% in severely ill patients^2,3^. The duration of delirium has consistently proven an independent predictor of longer hospitalization, higher mortality, and higher cost^4–6^, and delirium is often inadequately managed^7^. Antipsychotics drugs (APs) are usually selected for the pharmacological management of delirium^8^, and confusion and behavioural disturbances associated with hospitalization of elderly patients^9,10^.

COVID-19 patients are being treated with Chloroquine, Hydroxychloroquine^11,12^, Azithromycin,^13^ and Lopinavir/ritonavir^14^, for the main symptoms, with risk of QT prolongation and TdP, and others drug-drug interactions (DDIs) in coadministration with antipsychotics. The very ill patients being treated, are the most likely to suffer from QT prolongation, because of the clinical condition known to prolong QT, and the concomitant drugs, and clinicians need to mitigate that risk^15^. This use is complicated by age-related decline in drug metabolism^16^, high incidence of concomitant physical illnesses, drug-drug interactions, and heightened sensitivity to APs^17–19^.

In addition, APs are associated with a proarrhythmic state and an increased risk of sudden cardiac death (SCD), with no substantial differences between first and second generation antipsychotics, and a dose-response effect^20,21^. Antipsychotics (exception Aripiprazole and Lurasidone) seem to be associated with a prolonged QT interval and an increased risk of SCD^22^. There are differences among APs in the degree of cardiotoxic effects^23^. SCD describes the unexpected natural death from a cardiac cause, within a short a time of a person who often has no prior cardiological condition that would appear fatal^24^. Many antipsychotics show a certain degree of blockade of potassium channels coded by the human ether-à-go-go related gene (hERG), thus inducing a QT interval prolongation and increasing the risk of polymorphic ventricular tachycardia/ torsade de Pointes (TdP)^25,26^. The most common acquired cause of long QT syndrome and TdP is drug induced QT interval prolongation. Intensive Care Unit patients are particularly prone to experience a QTc interval prolongation mainly due to certain drugs that can prolong the repolarization phase, either by their action mechanism or through the interaction with other drugs^27^. Elderly patients are in particularly prone to proarrhythmic risk with antipsychotics^28^. Also, the potential for pharmacokinetic DDIs of APs are mostly mediated by cytochrome P450 (CYP) enzymes metabolism, therefore physicians should be aware of coadministered drugs that may inhibit or induce these CYP enzymes^29–31^.

The aim of our review is to analyze the risks of DDIs and cardiovascular adverse events of antipsychotics with the main treatments currently used for COVID-19. More so, SARS-CoV-2 primarily affects the elderly with other associated cardiovascular risk factors, which often require hospitalization, and treatment of which is esencial to knowing the risk and safety of antipsychotics associated with COVID-19 treatments.

## METHODS

Drugs currently used in the treatment of SARS-CoV-2^32^, chloroquine, hydroxychloroquine, lopinavir/ritonavir, remdesivir, and tocilizumab have been reviewed. Also azithromycin with promising results^13^, and potential agents therapeutic agents such as favipiravir^33^, baricitinib^34^, and anakinra^35^.

### Drug-drug interactions: antipsychotics and COVID19 drugs

Three databases were searched: (a) Lexicomp® Drug Interactions^36^, (b) Micromedex® Solutions Drugs Interactions^37^, and (c) Liverpool © Drug Interaction Group for COVID-19 therapies^38^. After an exhaustive review of various drugs interaction databases, the first two met the minimum quality criteria^39^, with good scores for scope, completeness, and ease of use^40^. The following information was extracted (when available): type of interaction, risk rating, and severity (both, according to the stratification in each database) and patient management recommendations.

### QT prolongation and TdP risk

To find out more about QT prolongation and TdP, the CredibleMeds® QTDrugs List^41^ was searched. CredibleMeds® classifies drugs based on their risk of QT prolongation or TdP. Risk category: (a) Known Risk of TdP, (b) Possible Risk of TdP, (c) Conditional Risk of TdP.

### Consensus recommendation

After the above information had been collected, the authors made consensus recommendations based on the following:

1. Not recommended (red zone), if (1) coadministration contraindicated in one or more databases, or (2) both drugs classified as “Known Risk of TdP” (risk of QT prolongation or TdP).
2. Recommended with caution. Two categories:

a. Potential interaction, which may require a dose adjustment, close monitoring, or choosing alternative agents, in two or more databases (orange zone).
b. b) Potential interaction intensity likely to be weak. Additional action/monitoring or dose adjustment unlikely to be required, in two or more databases (yellow zone).
3. Recommended (green zone), if little or no evidence of clinically significant interaction is expected in two or more databases.

When there were differences between databases, and it was not possible to reach an agreement by consensus, our recommendations reflect the most conservative approach.

When QT-prolongation is the main risk, we present the APs in two groups according to CredibleMeds® (“Possible” or “Conditional” risk) to increase the clinical information.

### Systematic review

Additionally, a systematic review was conducted according to PRISMA guidelines^42^ (registration number in PROSPERO CRD42020183202) to find clinical outcomes of drug-drug interactions between COVID-19 treatment and antipsychotics.

We researched the following databases: MEDLINE, EMBASE, and Web of Science. The search was restricted to the English language using database filters. No dates restriction were imposed for the updated search. The search strategy will follow syntax: antipsychotic* AND (chloroquine OR hydroxychloroquine OR lopinavir OR ritonavir OR remdesivir OR tocilizumab OR azithromycin OR favipiravir OR baricitinib OR anakinra). The reference list of all identified articles identified were examined for any additional studies nor found in the first search.

Inclusion and exclusion criteria: The characteristics for a study to be eligible were (1) Publication reporting clinically significant drug-drug interactions (between antipsychotics and COVID-10 treatment in adult patients (whatever the disease prescribed for), (2) Type of publication: randomized controlled trial, other controlled study, observational study, case report and case series. Exclusion criteria: (1) Only pharmacokinetic studies (no clinical outcome), (2) Unpublished studies or gray literature and conference articles.

Three reviewers (G.R., A.R., M.R.) independently screened titles for the research question and criteria. When articles were deemed to meet the inclusion criteria by either reviewer, the abstract was analyzed. Full texts were retrieved when the reviewers were in agreement that the article met the inclusion criteria. Disagreements were solved by a fourth independent reviewer (B.P.). Rayyan QCR was used for duplicated data and screening^43^.

Data extraction: Two authors reached an agreement regarding the type of data to be collected, and relevant information such as the study design, context of drug interactions, clinical outcome, other comedication data and the numbers and types of patients was recorded.

Risk of bias and generalizability concerns in comparative studies were evaluated with an adaptation of the Newcastle-Ottawa quality assessment scale for case control and cohort studies^44^, an eight-item tool designed to rate methodological aspects of case-control and cohort studies, which includes three domains: study selection, comparability of cohorts on the basis of the design or analysis, and exposure. The overall score ranges from 0 to 9. Risk of bias and generalizability concerns in case reports and case series were evaluated with the Drug Interaction Probability Scale (DIPS) tool to assess the quality of papers retained^45^, This 10-question tool evaluates drug interaction causation. Scores < 2 indicate doubtful drug interaction, scores 2–4 possible, 5–8 probable and scores ≥ 9 indicate highly probable drug interaction.

## RESULTS

The authors’ consensus recommendations are summarized in **Table 1**.

**Table 1.**
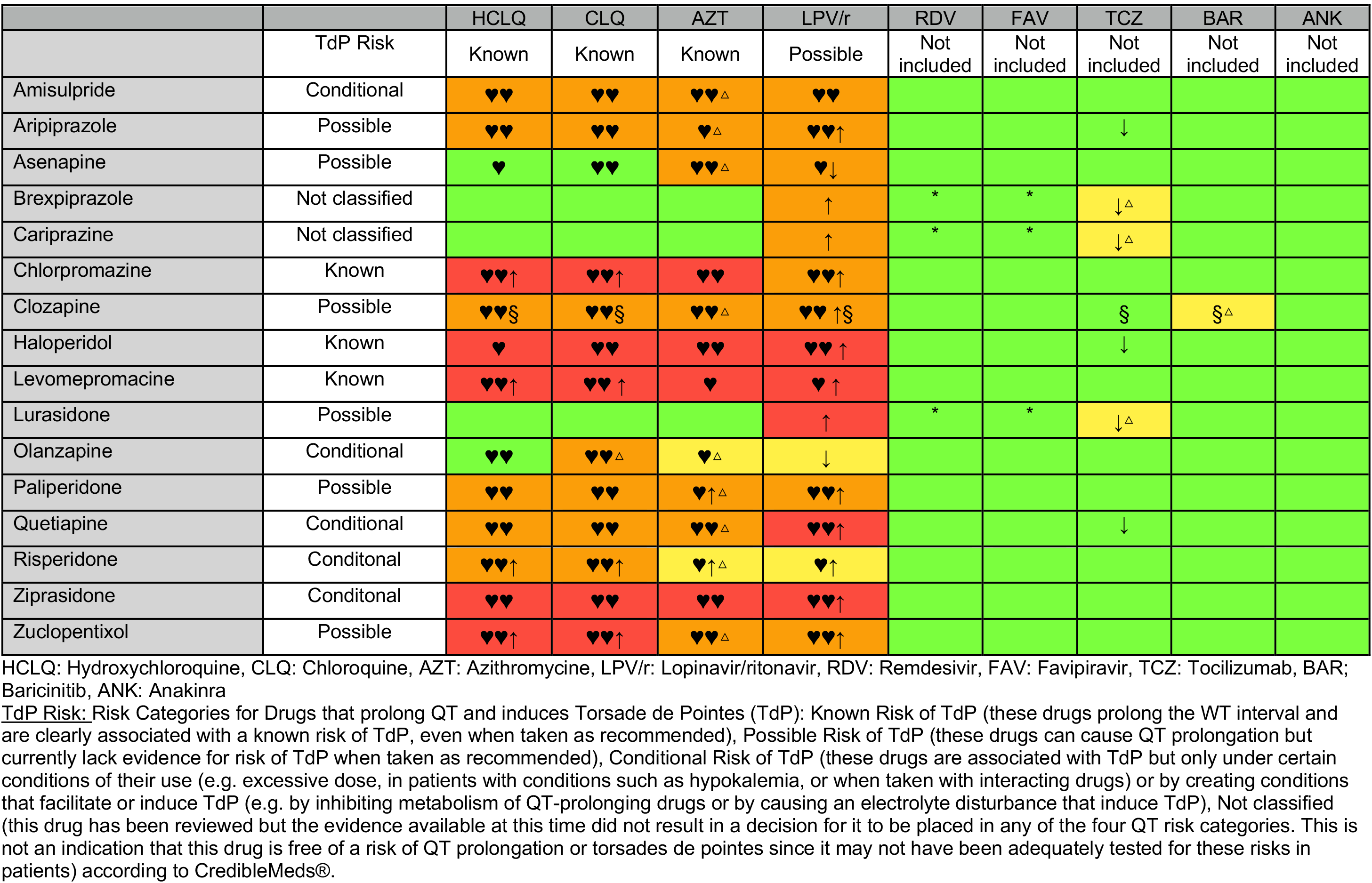
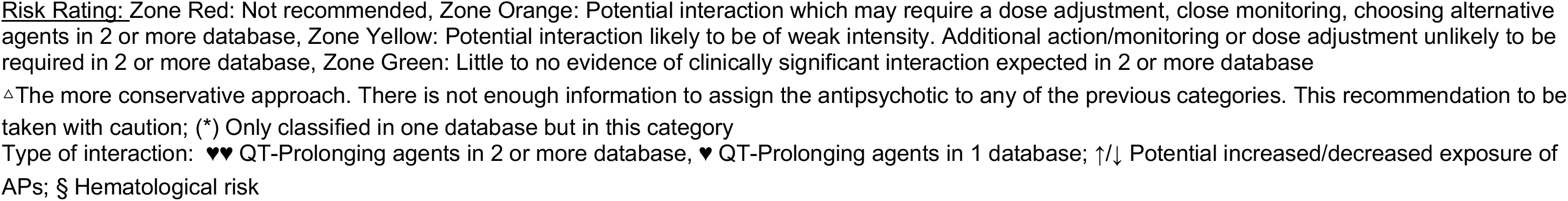
Author’s recommendation summary.

|  |  | HCLQ | CLQ | AZT | LPV/r | RDV | FAV | TCZ | BAR | ANK |
| --- | --- | --- | --- | --- | --- | --- | --- | --- | --- | --- |
|  | TdP Risk | Known | Known | Known | Possible | Not included | Not included | Not included | Not included | Not included |
| Amisulpride | Conditional | ♥♥ | ♥♥ | ♥♥△ | ♥♥ |  |  |  |  |  |
| Aripiprazole | Possible | ♥♥ | ♥♥ | ♥△ | ♥♥↑ |  |  | ↓ |  |  |
| Asenapine | Possible | ♥ | ♥♥ | ♥♥△ | ♥↓ |  |  |  |  |  |
| Brexpiprazole | Not classified |  |  |  | ↑ | * | * | ↓△ |  |  |
| Cariprazine | Not classified |  |  |  | ↑ | * | * | ↓△ |  |  |
| Chlorpromazine | Known | ♥♥↑ | ♥♥↑ | ♥♥ | ♥♥↑ |  |  |  |  |  |
| Clozapine | Possible | ♥♥§ | ♥♥§ | ♥♥△ | ♥♥↑§ |  |  | § | §△ |  |
| Haloperidol | Known | ♥ | ♥♥ | ♥♥ | ♥♥↑ |  |  | ↓ |  |  |
| Levomepromazine | Known | ♥♥↑ | ♥♥↑ | ♥ | ♥↑ |  |  |  |  |  |
| Lurasidone | Possible |  |  |  | ↑ | * | * | ↓△ |  |  |
| Olanzapine | Conditional | ♥♥ | ♥♥△ | ♥△ | ↓ |  |  |  |  |  |
| Paliperidone | Possible | ♥♥ | ♥♥ | ♥↑△ | ♥♥↑ |  |  |  |  |  |
| Quetiapine | Conditional | ♥♥ | ♥♥ | ♥♥△ | ♥♥↑ |  |  | ↓ |  |  |
| Risperidone | Conditonal | ♥♥↑ | ♥♥↑ | ♥↑△ | ♥↑ |  |  |  |  |  |
| Ziprasidone | Conditonal | ♥♥ | ♥♥ | ♥♥ | ♥♥↑ |  |  |  |  |  |
| Zuclopentixol | Possible | ♥♥↑ | ♥♥↑ | ♥♥△ | ♥♥↑ |  |  |  |  |  |

HCLQ: Hydroxychloroquine, CLQ: Chloroquine, AZT: Azithromycine, LPV/r: Lopinavir/ritonavir, RDV: Remdesivir, FAV: Favipiravir, TCZ: Tocilizumab, BAR: Baricitinib, ANK: Anakinra
TdP Risk: Risk Categories for Drugs that prolong QT and induces Torsade de Pointes (TdP): Known Risk of TdP (these drugs prolong the QT interval and are clearly associated with a known risk of TdP, even when taken as recommended), Possible Risk of TdP (these drugs can cause QT prolongation but currently lack evidence for risk of TdP when taken as recommended), Conditional Risk of TdP (these drugs are associated with TdP but only under certain conditions of their use (e.g. excessive dose, in patients with conditions such as hypokalemia, or when taken with interacting drugs) or by creating conditions that facilitate or induce TdP (e.g. by inhibiting metabolism of QT-prolonging drugs or by causing an electrolyte disturbance that induce TdP), Not classified (this drug has been reviewed but the evidence available at this time did not result in a decision for it to be placed in any of the four QT risk categories. This is not an indication that this drug is free of a risk of QT prolongation or torsades de pointes since it may not have been adequately tested for these risks in patients) according to CredibleMeds®.
Risk Rating: Zone Red: Not recommended, Zone Orange: Potential interaction which may require a dose adjustment, close monitoring, choosing alternative agents in 2 or more database, Zone Yellow: Potential interaction likely to be of weak intensity. Additional action/monitoring or dose adjustment unlikely to be required in 2 or more database, Zone Green: Little to no evidence of clinically significant interaction expected in 2 or more database
△The more conservative approach. There is not enough information to assign the antipsychotic to any of the previous categories. This recommendation to be taken with caution; (\*) Only classified in one database but in this category
Type of interaction: ♥♥ QT-Prolonging agents in 2 or more database, ♥ QT-Prolonging agents in 1 database; ↑/↓ Potential increased/decreased exposure of APs; § Hematological risk

### Drug-drug interaction between APs and Chloroquine/Hydroxychloroquine/Azithromycin

The primary interaction is the risk of QT prolongation and TdP because the three drugs for COVID-19, have “Known Risk of TdP” according to CredibleMeds®.

Recommended use of APs with these treatments is summarized in **Table 2**. Azithromycin is not included in the Liverpool© Drug Interaction Group, and therefore, some APs are marked as authors’ consensus agreement not possible. In that case, the recommendations reflect the most conservative approach, and they must be used with caution.

**Table 2.**
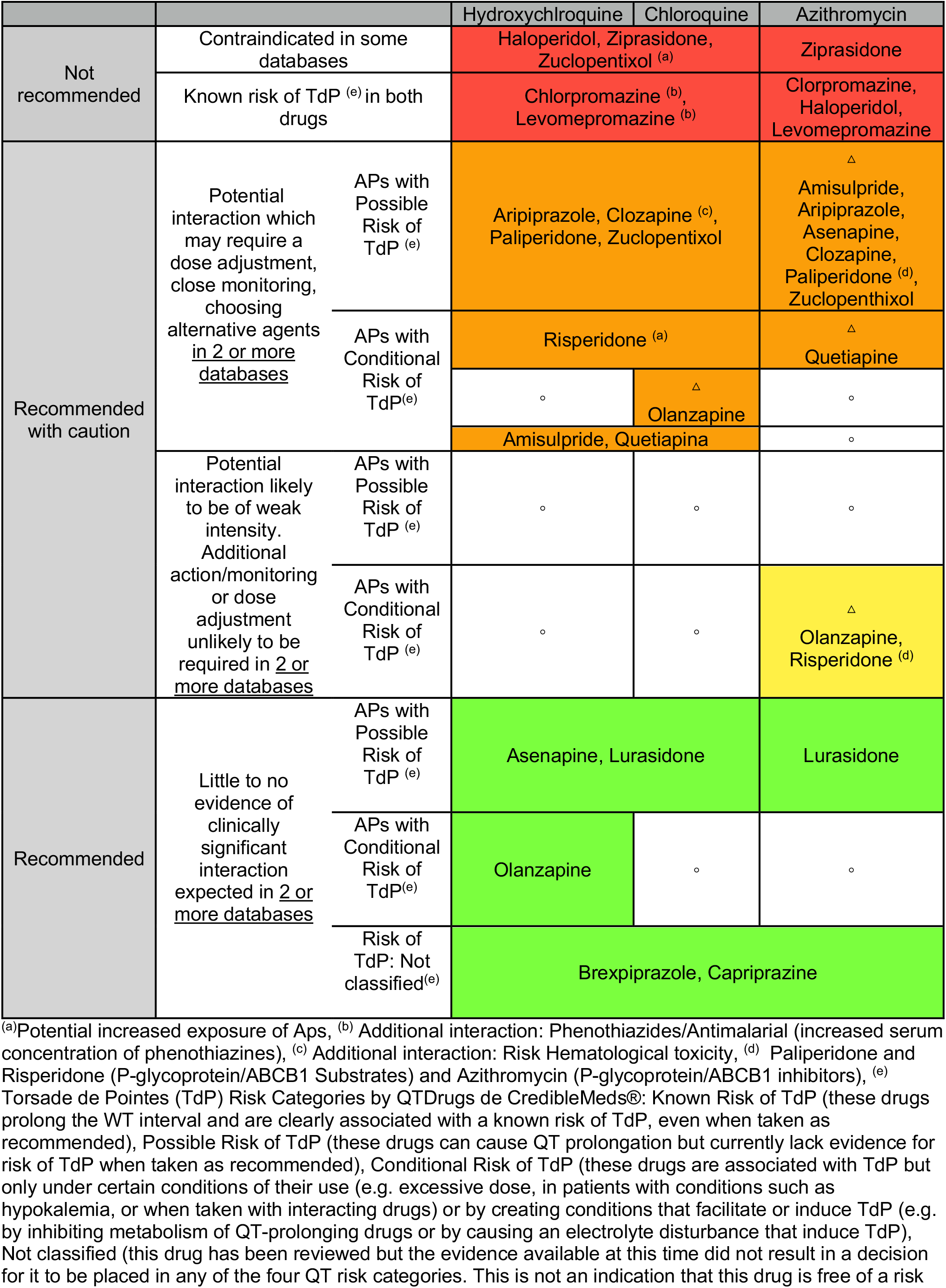
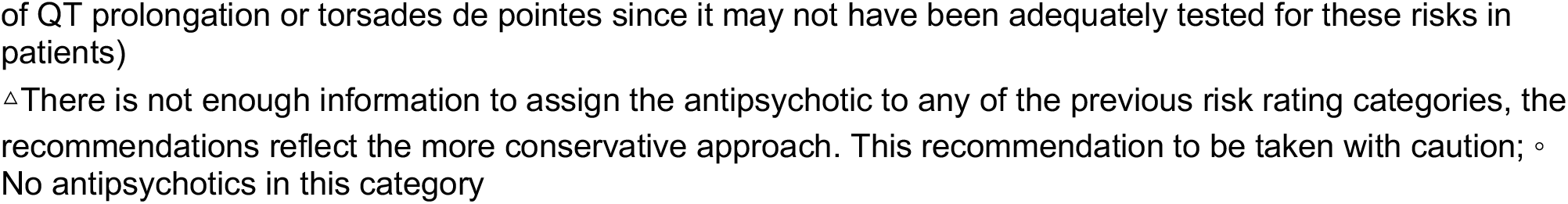
Drug-drug interaction between: Antipsychotics and Chloroquine/Hydroxychloroquine/Azithromycin.

The summary information about the risk rating, severity and patient management for each database is available in Supplement Tables 1–3.

**Table 3.**
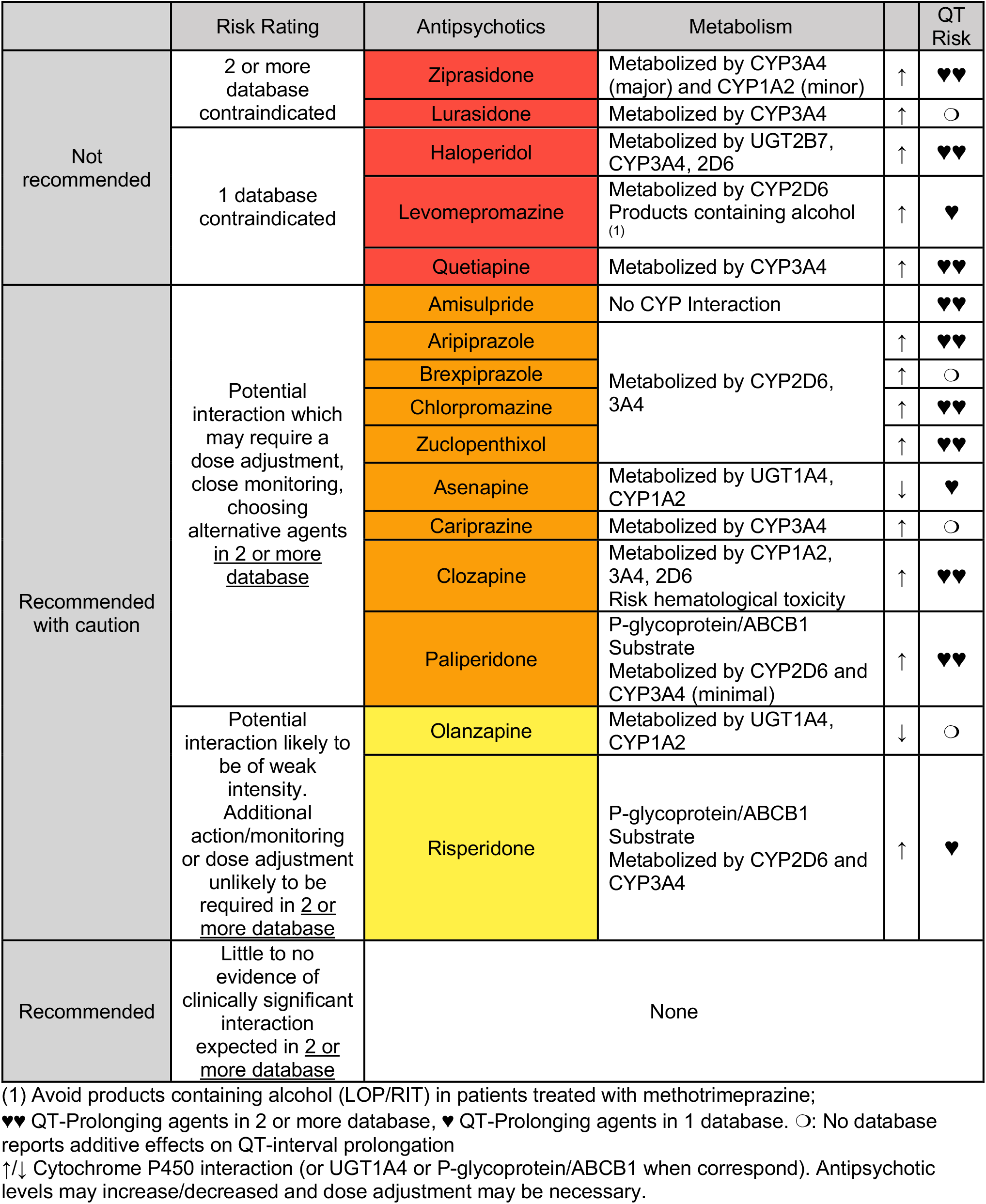
Drug-Drug interaction between Antipsychotics and Lopinavir/Ritonavir.

The recommended concomitant APs for use with these 3 COVID-19 drugs are: cariprazine, brexpiprazole and lurasidone. Asenapine may be used in combination with chloroquine and hydroxychloroquine, and olanzapine with hydroxychloroquine.

### Drug-drug interaction between APs and Lopinavir/Ritonavir

This coadministration involves CYP interactions and the risk of QT prolongation and TdP. Lopinavir/ritonavir have “Possible Risk of TdP” according to CredibleMeds®. Lopinavir is metabolized primarily by hepatic CYP3A4 isoenzymes, and ritonavir is known to inhibit CYP3A4, CYP2D6, and to a lesser extent CYP2C9/10 and CYP2C19. Lopinavir/ritonavir is a strong inhibitor of CYP3A4, less of CYP2D6 and P-Glycoprotein (P-gp), and is a CYP1A2 (weak) and UGT1A4 inducer.

**Table 3** summarizes the recommended uses of APs with lopinavir/ritonavir. There are no APs with little to no evidence of clinically significant interaction expected. We recommend olanzapine, with caution and monitoring patients for reduced clinical effect (ritonavir induction of CYP1A2), and risperidone, monitoring for increased AP effects and the risk of QT prolongation and/or TdP.

Information about risk ratings, severity and patient management in each database is summarized in Supplement Table 4.

### Drug-drug interaction between APs and Remdesivir, and Favipiravir

Lexicomp® Drug Interactions and Liverpool© Drug Interaction Group report that there are no significant risks. Remdesivir and Favipiravir are not included in the Micromedex® Solutions Drugs Interactions database.

### Drug-drug interaction between APs and Tocilizumab

Micromedex® Solutions Drug Interactions presents no drug-drug interaction, and although the Liverpool© Drug Interaction Group reports risk of hematological toxicity with clozapine, it does not present evidence of clinically significant risk with the rest of antipsychotics. However, according to Lexicomp® Drug Interactions, tocilizumab may decrease the serum concentration of CYP3A4 substrates like aripiprazole, brexpiprazole, cariprazine, haloperidol, lurasidone and quetiapine.

The information on risk rating, severity and patient management for each database is summarized in Supplement Table 5.

The authors’ consensus recommendation (see **Table 1**) is that brexpiprazole, cariprazine and lurasidone be used with caution. This is the most conservative approach, because they are not included in the Liverpool© database, and therefore, agreement is not possible.

### Drug-drug interaction between APs and Baricinitib

Baricinitib is not included in the Liverpool© Drug Interaction Group and Micromedex® Solutions Drugs Interactions shows no drug-drug interaction with antipsychotics. The summary information about the risk rating, severity and patient management for each database is available in eTable 6 in the Supplement. All antipsychotics are recommended, but clozapine because of hematological risk (risk rating in zone yellow).

### Drug-drug interaction between APs and Anakinra

Anakinra is not included in the Liverpool© Drug Interaction Group. Micromedex® Solutions Drugs Interactions and Lexicomp® Drug Interactions do not report any drug-drug interaction with antipsychotics.

### Results of the systematic review

Of the 391 studies were screened and assessed for eligibility. After applying inclusion and exclusion criteria (Supplement eFigure 1), only 7 case reports were selected for inclusion. Data from all included studies are summarized in **Table 4**. Two case reports are considered highly probable drug interaction according to DIPS, one with lopinavir/ritonavir and quetiapine (with priapism as the clinical outcome), and with ritonavir/indinavir and risperidone (with acute dystonia and tremor exacerbation as the clinical outcome). There were no deaths among the 8 patients, but two fatal outcome, one patient in coma (Risperidone+Ritonavir) and 2 others with Neuroleptic Malignant Syndrome (Ritonavir+Aripiprazole, and Ritonavir+Risperidone), however all of them remitted.

**Table 4.** Characteristics of the included studies involving patients with drug-drug interactions.

| Author/year | Title | Diagnosis | N | Treatment | Type of interaction | Clinical Outcome | Patient management | Drug Interaction Probability Scale (DIPS) |
| --- | --- | --- | --- | --- | --- | --- | --- | --- |
| Whiskey et al, 2018 <sup>61</sup> | Clozapine, HIV and neutropenia: a case report | Schizophrenia and HIV | 1 | CLZ 450 mg/day<br>RIT 100 mg/day<br>VAL 500mg/day<br>LMV and ZDV (not dose data) | Interaction CYP 3A4, CYP 2D6, CYP 1A2 and CYP 2C19 | Neutropenia attributable to the action and/or interaction of drugs. Unable to conclude whether due to the study variable (CLZ and RIT). | Evaluated the possibility of using GCSF with CLZ<br>VAL removal<br>Changed antiretrovirals (truvada and darunavir) and kept RIT | Possible |
| Geraci et al, 2010 <sup>62</sup> | Antipsychotic-induced priapism in an HIV patient: a cytochrome P450-mediated drug interaction | HIV | 1 | LPV 200 mg/RIT 50 mg/day<br>QUE 300mg/day | Interaction CYP 2D6 and CYP 3A4 | Priapism<br>Increased levels of psychotropic drugs | Anesthetized locally with 1% lidocaine and epinephrine and irrigated with dilute ephedrine and saline. | Highly probable |
| Aung et al., 2010 <sup>63</sup> | Increased Aripiprazole Concentrations in an HIV-Positive Male Concurrently Taking Duloxetine, Darunavir, and Ritonavir | HIV and Anxiety-Depressive Syndrome | 1 | ARI 50mg/day<br>RIT 100 mg/day | Interaction CYP 2D6 and CYP 3A4 | NMS<br>Increased levels of antipsychotic drugs | Disruption of the neuroleptics<br>Intensive fluid therapy administration | Probable |
| Pollack et al, 2009 <sup>64</sup> | Clinically significant adverse events from a drug interaction between quetiapine and atazanavir-ritonavir in two patients | First patient: HIV +BD<br>Second patient: HIV, anxiety disorder and | 2 | QUE400/day<br>RIT100/day | Interaction CYP 3A4 | Patient 1: Increase of 22 kg in 6 months.<br>Glucose level: 259mg/dL | Patient 1: RIT was replaced with Abacavir and the patient lost 5 kilograms per month and his serum glucose | Probable |
|  |  | substance abuse disorder |  |  |  | Patient 2: Delirium | level had normalized.<br>Patient 2: QUE stopped and resolution of mental status |  |
| Jover et al., 2002 <sup>65</sup> | Reversible Coma Caused By Risperidone-Ritonavir Interaction | HIV and manic episode | 1 | RIS 3 mg/day<br>RIT200 mg/day | Interaction CYP 2D6 and CYP 3A4 | Coma | APs stopped 24h later, neurological status returned to baseline and manic symptoms gradually reappeared | Probable |
| Kelly et al., 2002 <sup>66</sup> | Extrapyramidal Symptoms with Ritonavir/Indinavir plus Risperidone | HIV and Tourette's Syndrome | 1 | RIS 4 mg/day<br>RIT 400 mg /day | Interaction of both drugs via CYP 2D6 | Acute dystonia and tremor exacerbation | APs stopped 3 days later the symptoms improved significantly | Highly probable |
| Lee et al., 2000 <sup>67</sup> | Neuroleptic Malignant Syndrome associated with Use of Risperidone, Ritonavir, and Indinavir: A Case Report | HIV and acute psychotic episode | 1 | RIS 1,5 mg/day<br>RIT 400 mg/day | Interaction Track CYP 2D6 and CYP 3A4 | NMS | RIS and RIT were discontinued. Dantrolene and Bromocriptine were started. Clinical remission two weeks later | Probable |

ARI: Aripiprazol; BD: Bipolar Disorder; CLZ: Clozapine; GCSF: Granulocyte stimulating factor; LMV: Lamivudine; LPV: Lopinavir; NMS: Neuroleptic Malignant Syndrome; QUE: Quetiapine; RIT: Ritonavir; RIS: Risperidone; VAL: Valproic Acid; ZDV: Zidovudine

There was no study or case report about QT prolongation and/or TdP between the drug of our study.

## DISCUSSION

According to the DDIs databases and our systematic review, remdesivir, favipiravir, baricinitib, and anakinra may be concomitantly used with antipsychotics by clinicians for managing delirium, agitation or behavioral problems in COVID-19 patients with no risk of drug-drug interaction (except for hematological risk with clozapine and baricinitib reported only in one database).

Tocilizumab is quite safe for use in combination with antipsychotics, however, care should be taken with aripirazole, brexpiprazole, cariprazine, haloperidol, lurasidone and quetiapine, because of decreased exposure of APs, and with clozapine because of hematological risk). Tocilizumab binds to and inhibits the proinflammatory cytokine interleukin-6 (IL-6), and IL-6 decreased CYP3A4 mRNA by over 90%^46^.

Tocilizumab may restore CYP3A4 activity and thus increase CYP3A4 substrate metabolism. This effect may persist for several weeks following discontinuation of therapy due to the long half-life of tocilizumab. The COVID-19 treatments with the most risk for co-administration with antipsychotics are chloroquine, hydroxychloroquine, azithromycin, and lopinavir/ritonavir.

Most of the DDIs databases, point out the increased risk of prolonged QT interval with coadministration of chloroquine, hydroxychloroquine, and azithromycin with antipsychotics. Nevertheless, the cardiotoxicity of antimalarial drugs, with the exception of quinidine and halofantrine, which produce clinically significant prolongation of ventricular repolarization, remains to be confirmed^47,48^. In the present COVID scenario, Borba et al^49^ do not recommend the highest chloroquine dosage for patients critically ill with COVID-19 because of its potential safety hazards, especially when taken concurrently with azithromycin and oseltamivir. Risk of prolonged QTc (over 500 milliseconds) was higher in patients receiving high doses (600 mg twice daily for 10 days) than a low-dosage group (450 mg twice on Day 1 followed by once daily for four days), 18.9% versus 11.1%, respectively. It is worth mentioning that all patients were also receiving azithromycin, and nearly all were receiving oseltamivir (both also prolong QT interval)^50^. Although treatment with hydroxychloroquine, azithromycin, or both, was not compared with either treatment, in 1438 hospitalized patients with COVID-19, it was not significantly associated with differences in in-hospital mortality^51^. In addition, grading antipsychotics by torsadogenic risk is not a simple issue, and attributable risk varies depending on the source.^23,52^

Our work also suggests that the coadministration of protease inhibitors, such as lopinavir/ritonavir, with APs should be cataloged as the most problematic combination, because of the CYP DDIs^53^ and the risk of QT prolongation and/or TdP. Although none APs seem to be recommended for used in combination with lopinavir/ritonavir, olanzapine might be considered as the safest option according to DDIs databases and also because none case report describing DDI has been reported. Ritonavir decreases systemic exposure to olanzapine through the induction of both CYP1A2 and UGT. Thus, patients on concomitant olanzapine and ritonavir may require higher doses (up to 50% more) of olanzapine to achieve similar plasma concentrations^54,55^

### Strengths & Limitations

The use of drug interaction databases can be a useful clinical tool, but the lack of agreement between them limits their use^56–60^. Our study reviewed DDIs in 3 databases which fulfill quality criteria^39,40^, and a systematic review of the literature was also dome in search of possible clinical outcomes of these DDIs.

The main treatments used internationally for the treatment of COVID-19 were included, as well as most antipsychotics.

As limitations of our study, it should be noted that no studies were found beyond clinical cases which examined clinical results of the interaction between the drugs studied, so this information could not be presented.

## CONCLUSIONS

In conclusion, the growing use of antipsychotics in COVID19 patients calls on the urges development evidence-based guidelines that can help clinicians decide on the safest treatment combination and the level of monitoring that would be warranted for each particular patient. The selection of concomitant COVID-19 medications and antipsychotics should be evidence-based and closely monitored. The main interactions between COVID-19 drugs and antipsychotics are the risk of QT prolongation and/or TdP, and CYP interactions.

## Data Availability

Data is available in the article and supplementary material

## ACKNOWLEDGEMENT SECTION

We would like to thank Carmen Jimenez, who encouraged us to start this work.

## REFERENCES

1. National Institute for Health and Care Excellence (NICE) in collaboration with NHS England and NHS Improvement. Managing COVID-19 symptoms (including at the end of life) in the community: summary of NICE guidelines. BMJ. 2020;369:m1461. doi:10.1136/bmj.m1461

2. Mao L, Jin H, Wang M, et al. Neurologic Manifestations of Hospitalized Patients With Coronavirus Disease 2019 in Wuhan, China. JAMA Neurol. Published online April 10, 2020. doi:10.1001/jamaneurol.2020.1127

3. Helms J, Kremer S, Merdji H, et al. Neurologic Features in Severe SARS-CoV-2 Infection. N Engl J Med. 2020;0(0):null. doi:10.1056/NEJMc2008597

4. Ely E, Gautam S, Margolin R, et al. The impact of delirium in the intensive care unit on hospital length of stay. Intensive Care Med. 2001;27(12):1892–1900. doi:10.1007/s00134-001-1132-2

5. Ely EW, Shintani A, Truman B, et al. Delirium as a predictor of mortality in mechanically ventilated patients in the intensive care unit. JAMA. 2004;291(14):1753–1762. doi:10.1001/jama.291.14.1753

6. Vasilevskis EE, Chandrasekhar R, Holtze CH, et al. The Cost of ICU Delirium and Coma in the Intensive Care Unit Patient. Med Care. 2018;56(10):890–897. doi:10.1097/MLR.0000000000000975

7. O’Hanlon S, Inouye SK. Delirium: a missing piece in the COVID-19 pandemic puzzle. Age Ageing. Published online May 6, 2020. doi:10.1093/ageing/afaa094

8. Kotfis K, Marra A, Ely EW. ICU delirium – a diagnostic and therapeutic challenge in the intensive care unit. Anaesthesiol Intensive Ther. 2018;50(2):160–167. doi:10.5603/AIT.a2018.0011

9. Jin B, Liu H. Comparative efficacy and safety of therapy for the behavioral and psychological symptoms of dementia: a systemic review and Bayesian network meta-analysis. J Neurol. 2019;266(10):2363–2375. doi:10.1007/s00415-019-09200-8

10. van der Linde RM, Dening T, Matthews FE, Brayne C. Grouping of behavioural and psychological symptoms of dementia. Int J Geriatr Psychiatry. 2014;29(6):562–568. doi:10.1002/gps.4037

11. Colson P, Rolain J-M, Lagier J-C, Brouqui P, Raoult D. Chloroquine and hydroxychloroquine as available weapons to fight COVID-19. Int J Antimicrob Agents. Published online March 4, 2020:105932. doi:10.1016/j.ijantimicag.2020.105932

12. Shukla AM, Archibald LK, Shukla AW, Mehta HJ, Cherabuddi K. Chloroquine and hydroxychloroquine in the context of COVID-19. Drugs Context. 2020;9. doi:10.7573/dic.2020-4-5

13. Gautret P, Lagier J-C, Parola P, et al. Hydroxychloroquine and azithromycin as a treatment of COVID-19: results of an open-label non-randomized clinical trial. Int J Antimicrob Agents. Published online ,March 20, 2020:105949. doi:10.1016/j.ijantimicag.2020.105949

14. Cao B, Wang Y, Wen D, et al. A Trial of Lopinavir-Ritonavir in Adults Hospitalized with Severe Covid-19. N Engl J Med. Published online March 18, 2020. doi:10.1056/NEJMoa2001282

15. Giudicessi JR, Noseworthy PA, Friedman PA, Ackerman MJ. Urgent Guidance for Navigating and Circumventing the QTc-Prolonging and Torsadogenic Potential of Possible Pharmacotherapies for Coronavirus Disease 19 (COVID-19. Mayo Clin Proc. 2020;0(0). doi:10.1016/j.mayocp.2020.03.024

16. Zanger UM, Schwab M. Cytochrome P450 enzymes in drug metabolism: regulation of gene expression, enzyme activities, and impact of genetic variation. Pharmacol Ther. 2013;138(1):103–141. doi:10.1016/j.pharmthera.2012.12.007

17. Rivière J, van der Mast RC, Vandenberghe J, Van Den Eede F. Efficacy and Tolerability of Atypical Antipsychotics in the Treatment of Delirium: A Systematic Review of the Literature. Psychosomatics. 2019;60(1):18–26. doi:10.1016/j.psym.2018.05.011

18. Gareri P, De Fazio P, Manfredi VGL, De Sarro G. Use and safety of antipsychotics in behavioral disorders in elderly people with dementia. J Clin Psychopharmacol. 2014;34(1):109–123. doi:10.1097/JCP.0b013e3182a6096e

19. Maher AR, Maglione M, Bagley S, et al. Efficacy and comparative effectiveness of atypical antipsychotic medications for off-label uses in adults: a systematic review and meta-analysis. JAMA. 2011;306(12):1359–1369. doi:10.1001/jama.2011.1360

20. Ray WA, Chung CP, Murray KT, Hall K, Stein CM. Atypical antipsychotic drugs and the risk of sudden cardiac death. N Engl J Med. 2009;360(3):225–235. doi:10.1056/NEJMoa0806994

21. Salvo F, Pariente A, Shakir S, et al. Sudden cardiac and sudden unexpected death related to antipsychotics: A meta-analysis of observational studies. Clin Pharmacol Ther. 2016;99(3):306–314. doi:10.1002/cpt.250

22. Acciavatti T, Martinotti G, Corbo M, et al. Psychotropic drugs and ventricular repolarisation: The effects on QT interval, T-peak to T-end interval and QT dispersion. J Psychopharmacol (Oxf). 2017;31(4):453–460. doi:10.1177/0269881116684337

23. Leucht S, Cipriani A, Spineli L, et al. Comparative efficacy and tolerability of 15 antipsychotic drugs in schizophrenia: a multiple-treatments meta-analysis. Lancet Lond Engl. 2013;382(9896):951–962. doi:10.1016/S0140-6736(13)60733-3

24. Wellens HJJ, Schwartz PJ, Lindemans FW, et al. Risk stratification for sudden cardiac death: current status and challenges for the future. Eur Heart J. 2014;35(25):1642–1651. doi:10.1093/eurheartj/ehu176

25. Roden DM. Drug-induced prolongation of the QT interval. N Engl J Med. 2004;350(10):1013–1022. doi:10.1056/NEJMra032426

26. Testai L, Bianucci AM, Massarelli I, Breschi MC, Martinotti E, Calderone V. Torsadogenic cardiotoxicity of antipsychotic drugs: a structural feature, potentially involved in the interaction with cardiac HERG potassium channels. Curr Med Chem. 2004;11(20):2691–2706. doi:10.2174/0929867043364351

27. Etchegoyen CV, Keller GA, Mrad S, Cheng S, Di Girolamo G. Drug-induced QT Interval Prolongation in the Intensive Care Unit. Curr Clin Pharmacol. 2017;12(4):210–222. doi:10.2174/1574884713666180223123947

28. Vieweg WVR, Wood MA, Fernandez A, Beatty-Brooks M, Hasnain M, Pandurangi AK. Proarrhythmic risk with antipsychotic and antidepressant drugs: implications in the elderly. Drugs Aging. 2009;26(12):997–1012. doi:10.2165/11318880-000000000-00000

29. Kennedy WK, Jann MW, Kutscher EC. Clinically significant drug interactions with atypical antipsychotics. CNS Drugs. 2013;27(12):1021–1048. doi:10.1007/s40263-013-0114-6

30. Conley RR, Kelly DL. Drug-drug interactions associated with second-generation antipsychotics: considerations for clinicians and patients. Psychopharmacol Bull. 2007;40(1):77–97.

31. King JR, Wynn H, Brundage R, Acosta EP. Pharmacokinetic enhancement of protease inhibitor therapy. Clin Pharmacokinet. 2004;43(5):291–310. doi:10.2165/00003088-200443050-00003

32. Sanders JM, Monogue ML, Jodlowski TZ, Cutrell JB. Pharmacologic Treatments for Coronavirus Disease 2019 (COVID-19): A Review. JAMA. Published online April 13, 2020. doi:10.1001/jama.2020.6019

33. Jean S-S, Lee P-I, Hsueh P-R. Treatment options for COVID-19: The reality and challenges. J Microbiol Immunol Infect Wei Mian Yu Gan Ran Za Zhi. Published online April 4, 2020. doi:10.1016/j.jmii.2020.03.034

34. Cantini F, Niccoli L, Matarrese D, Nicastri E, Stobbione P, Goletti D. Baricitinib therapy in COVID-19: A pilot study on safety and clinical impact. J Infect. Published online April 22, 2020. doi:10.1016/j.jinf.2020.04.017

35. Monteagudo LA, Boothby A, Gertner E. Continuous Intravenous Anakinra Infusion to Calm the Cytokine Storm in Macrophage Activation Syndrome. ACR Open Rheumatol. Published online April 8, 2020. doi:10.1002/acr2.11135

36. Wolters Kluwer. Wolters Kluwer. Lexicomp® Interactions Module [Internet]. Accessed May 9, 2020. https://www.wolterskluwercdi.com/lexicomp-online/

37. Truven Health Analytics. Micromedex® Solutions Drugs Interaction [electronic version]. Accessed May 9, 2020. http://www.micromedexsolutions.com.

38. University of Liverpool. Liverpool COVID-19 Interactions. Accessed May 9, 2020. https://www.covid19-druginteractions.org/prescribing-resources

39. Rodríguez-Terol A, Caraballo MO, Palma D, et al. Calidad estructural de las bases de datos de interacciones. Farm Hosp. 2009;33(3):134–146. doi:10.1016/S1130-6343(09)71155-9

40. Patel RI, Beckett RD. Evaluation of resources for analyzing drug interactions. J Med Libr Assoc JMLA. 2016;104(4):290–295. doi:10.3163/1536-5050.104.4.007

41. Woosley, RL, Heise, CW Romero, KA. CredibleMeds ®. Accessed May 9, 2020. https://www.crediblemeds.org/

42. Moher D, Liberati A, Tetzlaff J, Altman DG, PRISMA Group. Preferred reporting items for systematic reviews and meta-analyses: the PRISMA statement. PLoS Med. 2009;6(7):e1000097. doi:10.1371/journal.pmed.1000097

43. Ouzzani M, Hammady H, Fedorowicz Z, Elmagarmid A. Rayyan-a web and mobile app for systematic reviews. Syst Rev. 2016;5(1):210. doi:10.1186/s13643-016-0384-4

44. Stang A. Critical evaluation of the Newcastle-Ottawa scale for the assessment of the quality of nonrandomized studies in meta-analyses. Eur J Epidemiol. 2010;25(9):603–605. doi:10.1007/s10654-010-9491-z

45. Horn JR, Hansten PD, Chan L-N. Proposal for a new tool to evaluate drug interaction cases. Ann Pharmacother. 2007;41(4):674–680. doi:10.1345/aph.1H423

46. Aitken AE, Morgan ET. Gene-specific effects of inflammatory cytokines on cytochrome P450 2C, 2B6 and 3A4 mRNA levels in human hepatocytes. Drug Metab Dispos Biol Fate Chem. 2007;35(9):1687–1693. doi:10.1124/dmd.107.015511

47. White NJ. Cardiotoxicity of antimalarial drugs. Lancet Infect Dis. 2007;7(8):549–558. doi:10.1016/S1473-3099(07)70187-1

48. Haeusler IL, Chan XHS, Guérin PJ, White NJ. The arrhythmogenic cardiotoxicity of the quinoline and structurally related antimalarial drugs: a systematic review. BMC Med. 2018;16(1):200. doi:10.1186/s12916-018-1188-2

49. Borba MGS, Val FFA, Sampaio VS, et al. Effect of High vs Low Doses of Chloroquine Diphosphate as Adjunctive Therapy for Patients Hospitalized With Severe Acute Respiratory Syndrome Coronavirus 2 (SARSCoV-2) Infection: A Randomized Clinical Trial. JAMA Netw Open. 2020;3(4):e208857. doi:10.1001/jamanetworkopen.2020.8857

50. Fihn SD, Perencevich E, Bradley SM. Caution Needed on the Use of Chloroquine and Hydroxychloroquine for Coronavirus Disease 2019. JAMA Netw Open. 2020;3(4):e209035–e209035. doi:10.1001/jamanetworkopen.2020.9035

51. Rosenberg ES, Dufort EM, Udo T, et al. Association of Treatment With Hydroxychloroquine or Azithromycin With In-Hospital Mortality in Patients With COVID-19 in New York State. JAMA. Published online May 11, 2020. doi:10.1001/jama.2020.8630

52. Raschi E, Poluzzi E, Godman B, et al. Torsadogenic risk of antipsychotics: combining adverse event reports with drug utilization data across Europe. PloS One. 2013;8(11):e81208. doi:10.1371/journal.pone.0081208

53. Ernest CS, Hall SD, Jones DR. Mechanism-based inactivation of CYP3A by HIV protease inhibitors. J Pharmacol Exp Ther. 2005;312(2):583–591. doi:10.1124/jpet.104.075416

54. Penzak SR, Hon YY, Lawhorn WD, Shirley KL, Spratlin V, Jann MW. Influence of ritonavir on olanzapine pharmacokinetics in healthy volunteers. J Clin Psychopharmacol. 2002;22(4):366–370. doi:10.1097/00004714-200208000-00006

55. Meemken L, Hanhoff N, Tseng A, et al. Drug-Drug Interactions With Antiviral Agents in People Who Inject Drugs Requiring Substitution Therapy. Ann Pharmacother. 2015;49(7):796–807.

56. Abarca J, Malone DC, Armstrong EP, et al. Concordance of severity ratings provided in four drug interaction compendia. J Am Pharm Assoc JAPhA. 2004;44(2):136–141. doi:10.1331/154434504773062582

57. Chao SD, Maibach HI. Lack of drug interaction conformity in commonly used drug compendia for selected at-risk dermatologic drugs. Am J Clin Dermatol. 2005;6(2):105–111. doi:10.2165/00128071-200506020-00005

58. Fulda TR, Valuck RJ, Zanden JV, Parker S, Byrns PJ, Acknowledgments UPDURAP of the UPDURAP are listed in the. Disagreement Among Drug Compendia on Inclusion and Ratings of Drug-Drug Interactions. Curr Ther Res. 2000;8(61):540–548.

59. Vitry AI. Comparative assessment of four drug interaction compendia. Br J Clin Pharmacol. 2007;63(6):709–714. doi:10.1111/j.1365-2125.2006.02809.x

60. Monteith S, Glenn T. A comparison of potential psychiatric drug interactions from six drug interaction database programs. Psychiatry Res. 2019;275:366–372. doi:10.1016/j.psychres.2019.03.041

61. Whiskey E, O’Flynn D, Taylor D. Clozapine, HIV and neutropenia: a case report. Ther Adv Psychopharmacol. 2018;8(12):365–369. doi:10.1177/2045125318804499

62. Geraci MJ, McCoy SL, Crum PM, Patel RA. Antipsychotic-induced priapism in an HIV patient: a cytochrome P450-mediated drug interaction. Int J Emerg Med. 2010;3(2):81–84. doi:10.1007/s12245-010-0175-y

63. Aung GL, O’Brien JG, Tien PG, Kawamoto LS. Increased aripiprazole concentrations in an HIV-positive male concurrently taking duloxetine, darunavir, and ritonavir. Ann Pharmacother. 2010;44(11):1850–1854. doi:10.1345/aph.1P139

64. Pollack TM, McCoy C, Stead W. Clinically significant adverse events from a drug interaction between quetiapine and atazanavir-ritonavir in two patients. Pharmacotherapy. 2009;29(11):1386–1391. doi:10.1592/phco.29.11.1386

65. Jover F, Cuadrado J-M, Andreu L, Merino J. Reversible coma caused by risperidone-ritonavir interaction. Clin Neuropharmacol. 2002;25(5):251–253. doi:10.1097/00002826-200209000-00004

66. Kelly DV, Béïque LC, Bowmer MI. Extrapyramidal symptoms with ritonavir/indinavir plus risperidone. Ann Pharmacother. 2002;36(5):827–830. doi:10.1345/aph.1A335

67. Lee SI, Klesmer J, Hirsch BE. Neuroleptic malignant syndrome associated with use of risperidone, ritonavir, and indinavir: a case report. Psychosomatics. 2000;41(5):453–454. doi:10.1176/appi.psy.41.5.453

